# Psychometric properties of the Sleep Hygiene Index in a large Italian community sample

**DOI:** 10.1101/2021.05.26.21257845

**Authors:** Andrea Zagaria, Andrea Ballesio, Alessandro Musetti, Vittorio Lenzo, Maria C. Quattropani, Lidia Borghi, Giorgia Margherita, Emanuela Saita, Gianluca Castelnuovo, Maria Filosa, Laura Palagini, Giuseppe Plazzi, Caterina Lombardo, Christian Franceschini

## Abstract

**Objective/Background:** Poor sleep hygiene is considered an exacerbating and perpetuating factor of sleep disturbances and is also associated with poor mental health. The Sleep Hygiene Index (SHI) is a self-report measure assessing adherence to sleep hygiene practices. The aim of this study was to estimate the psychometric properties of the SHI in an Italian representative sample of the general population, following a formative measurement approach.

**Patients/Methods:** Participants (n=6276; M=33.62, SD=13.45) completed the SHI alongside measures of sleep disturbance, depression, anxiety, and stress. To consider the item formative nature, sets of item-composites weighted by means of canonical correlation analysis was created and a confirmatory factor analysis (CFA) was implemented. Factorial invariance tests were computed considering both presence of sleep problems and presence of emotional distress symptoms as grouping variables.

**Results and Conclusions:** CFA confirmed the unidimensional structure of SHI. Internal consistency was acceptable (ω=0.752). Test-retest reliability at 8-10 months presented an ICC of 0.666. SHI significantly correlated with sleep, depression, anxiety and stress symptoms (r range from 0.358 to 0.500). Configural and metric invariance were reached for both grouping variables. Partial scalar invariance was obtained only across emotional distress groups. People with emotional symptoms reported higher latent means on the sleep hygiene dimension. Findings support the validity and reliability of the Italian version of the SHI. Importantly, the SHI showed robust psychometric properties both in healthy individuals and in individual reporting mental health symptoms. Thus, it is advisable to use this version of the SHI in both research and clinical practice.

## 1. Introduction

Sleep hygiene may be defined as a set of behavioural and environmental recommendations that are shown to facilitate or inhibit sleep in empirical research (e.g., Edinger et al. 2017). Although firstly mentioned in the pioneering work by Kleitman (1939), the concept of sleep hygiene gained considerable importance in the last three decades. Poor sleep hygiene is traditionally considered as a core maintaining factor of chronic insomnia in behavioural (e.g., Spielman et al. 1987) and cognitive models (e.g., Morin 1993). Additionally, poor sleep hygiene is involved in the maintenance of other sleep disorders including obstructive sleep apnoea (e.g., Jung et al. 2019), shift work disorder (Booker et al. 2020), and restless leg syndrome (Sönmez & Aksoy Derya, 2018). Conversely, the implementation of sleep hygiene practices may have the potential to address the public health concern of sleep complaints in the general population (Irish et al. 2015). Related to this, sleep hygiene has been found to be predictive of better mental health (e.g., lower depression and higher well-being) in healthy samples (Peach et al. 2016) and in individuals with depression (Gupta et al. 2019).

The adherence to sleep hygiene practices can be easily assessed using self-reported measures, such as the Sleep Hygiene Awareness and Practice Scale (SHAPS; Lacks & Rotert, 1986), the Sleep Hygiene Self-Test (SHST; Blake & Gòmez, 1998) and the Sleep Hygiene Index (SHI; Mastin, Bryson & Corwyn, 2006). While the first two tools have been found to have relatively low psychometric properties, the SHI seems to provide a valid and reliable measure of the sleep hygiene. Specifically, the SHI showed a good two-week test-retest reliability as well as strongly associations with sleep quality and with the associated features of inadequate sleep hygiene (Mastin, Bryson & Corwyn, 2006). In addition, in comparison to other tools, the SHI was initially developed following a clear rationale for the item selection, namely the diagnostic criteria for inadequate sleep hygiene proposed in the third edition of the International Classification of Sleep Disorders (American Academy of Sleep Medicine, 2014). The SHI has been adapted and validated in different populations and languages, including Turkish (Ozdemir et al., 2015), Spanish (Prados et al., 2021), Brazilian (Tonon et al., 2020), Chinese (Tang et al., 2021), Nigerian (Seun-Fadipe et al., 2018) and Arabic (Costa et al., 2021).

The aim of this study was to estimate the psychometric properties of the SHI in an Italian representative sample of the general population. More specifically, the objectives of this study were: 1) analysing the factor structure of the Italian adaptation of the SHI (Palagini & Manni, 2017) adopting a formative indicators approach as postulated by the original authors, as well as evaluate the adequacy of the factor model; 2) examining the reliability (i.e., internal consistency and test-retest reliability) and the predictive validity of the tool. Finally, to the best of our knowledge, no studies have explored the factorial invariance of the tool according to clinical variables. Thus, another objective was to evaluate latent mean differences on the sleep hygiene dimension across people with self-reported sleep problems (i.e., with or without sleep problems groups) and across people with self-reported emotional distress (i.e., with or without symptoms groups) by assessing the factorial invariance of the construct.

## 2. Material and Methods

### 2.1. Procedure and sampling

A cross-sectional design was used for the primary analysis. A subgroup of participants also completed the SHI in a follow-up in order to estimate test-retest reliability (see below). This study was conducted in accordance with the Declaration of Helsinki. Ethical clearance was obtained from the Ethics Committee of the Center for Research and Psychological Intervention (CERIP) of the University of Messina (n. 37442). An online survey using Microsoft Azure was used to collect all data. On the online platform, information and consent to the processing of personal data were requested. Anonymity was guaranteed for all participants involved in this study by the creation of an authorless identification code. Google Cloud platform was used to create the administered questionnaires. The study was publicised through numerous sources, including university communication platform, online forums and social networks. All data were collected during spring 2020. Specifically, the survey was available online from 10 March 2020 to 4 May 2020. A total of 6276 participants were recruited through announcements in five Italian universities (La Statale University of Milan, Catholic University of Milan, University of Parma, University of Messina, Federico II University of Naples) using a convenience sampling procedure. Only people who completed all questionnaires and who lived in Italy were included in our investigation.

### 2.2. Measures

#### 2.2.1. Demographic information

A sociodemographic questionnaire was created to obtain information about gender, age, region of residence, marital status, number of children, education level and occupation of all participants.

#### 2.2.2. Sleep Hygiene Index (SHI)

The SHI, originally developed by Mastin et coll. (2006), is a 13 item self-reported instrument implemented to assess practices and behaviours related to sleep hygiene. Participants are asked to rate the frequency to which they have engaged in specific behaviours on a 5-point Likert scale (“never, “rarely, “sometimes”, “frequently”, “always”). A total score is calculated by summing each SHI item, where higher scores are indicative of poor sleep hygiene. The global assessment score ranges from 13 to 65.

#### 2.2.3. Medical Outcome Study (MOS)

The MOS Sleep Scale (Hays & Stewart, 1992) is a self-reported instrument evaluating sleep quality and duration. It consists of 12 items reflecting six subscales: sleep disturbance, sleep adequacy, sleep quantity, snoring, awaken short of breath or with headache, and daytime somnolence. Two sleep problems composite index can also be computed by summing 6 or 9 items of the MOS. Except for sleep quantity that is scored as the number of hours of sleep reported, all subscales or composite scores are transformed on a 0–100 metric where higher scores indicate a greater occurrence of sleep symptoms. Satisfactory psychometric properties were found in the general population (e.g., Hays, Martin & Sesti, 2005).

#### 2.2.4. Depression Anxiety Stress Scale-21 (DASS-21)

The DASS-21 (Lovibond & Lovibond, 1995), is a short version of a self-reported questionnaire that assess emotional distress across three domains: depression, anxiety, and stress. Participants are asked to indicate the severity to which they have experienced each symptom in the previous week on a 4-point Likert scale. Each subscale contains 7 items, where higher scores indicate more severe emotional distress. Subscales scores are calculated by summing relevant item and are interpreted as normal, mild, moderate, severe, or extremely severe. A general distress score is also calculated by summing all DASS items. The validity and reliability of the DASS 21 questionnaire has been provided in the Italian population (Bottesi et al., 2015)

### 2.3. Statistical analyses

All data were analysed by means of IBM SPSS 23 and MPLUS 8.4. Descriptive statistics were used to describe SHI items and participants’ characteristics. Prior to analysis, deviations from univariate normality were analysed. Skewness and kurtosis values greater than |1| indicates a non-negligible violation of normality (Tabachnick & Fidell, 2007). As postulated by the original authors (Mastin et al., 2006), SHI was conceptualized following a formative indicators approach. In this perspective, indicators could be viewed as causing rather than caused by the latent variable (i.e., causal items vs reflective items) (MacCallum & Browne 1993). Causal items need a different set of assumptions and a different measurement model testing: exploratory factor analysis and confirmatory factor analysis assume that indicators are reflective indicators (Bollen & Diamantopoulos, 2017; Coltman et al., 2008). Treiblmaier et al., (2011) implemented a two-step procedure to test formative measurement model via common factors. First, SHI indicators were split into four sets in order to identify maximally correlated composites of indicators. Based on these sets, a non-linear canonical correlation analysis was implemented and optimal scoring weights (i.e., canonical coefficients) were obtained. By means of the canonical coefficients, we weighted the observed variables constituting each composite, and four item-composite scores were computed. Finally, we treated the weighted composites as reflective indicators of the sleep hygiene construct. This procedure allowed us to implement a traditional confirmatory factor analysis (CFA). To take into account the items non-normality, a robust maximum likelihood method of estimation was applied to test the hypothesized model. According to a multifaced conception of model fit (Tanaka, 1993), several goodness-of-fit indices with respective cut-off values were considered (Hu & Bentler, 1999): RMSEA (< 0.08 indicates acceptable fit); CFI (> 0.90 indicates acceptable fit); TLI (> 0.90 indicates acceptable fit); SRMR (< 0.08 indicates acceptable fit). Traditional χ2 statistics were reported but not considered due to its sensitivity to large sample size (Cheung & Rensvold, 2002). SHI reliability was assessed with McDonald’s omega including the weighted composites (McDonald, 1999). Omega coefficients was preferred to traditional alpha due to the strict assumption of the Cronbach’s coefficient (see in this regard Hermosilla & Alvarado, 2016). Value ≥.70 are considered adequate (Bagozzi & Yi, 2012). Test-retest reliability at 8-10 months was assessed on a subsample of 648 subjects using the intraclass correlation coefficient (ICC 2,1; see the convention of Shrout & Fleiss, 1979). Subsequently, given the high correlations between sleep hygiene and sleep and emotional symptoms (Peach et al. 2016; Edinger et al. 2017; Gupta et al. 2019), Pearson’s correlation coefficients between SHI, MOS, and DASS scores were computed in order to test the predictive validity of the tool. Correlation lower than .30 were considered small, between 0.30–0.49 moderate, and at least 0.50 strong (Cohen, 1988). Finally, factorial invariance tests were computed within the conceptual framework proposed by Meredith (1993) by means of multi-group confirmatory factor analysis (MG-CFA; Bollen, 1989). Testing for measurement invariance at the item-composites level of aggregation can be feasible (Little et al., 2013). Configural invariance, metric invariance and scalar invariance were tested. Given the sensibility of chi-square tests to large sample size, differences in alternative fit indices were used to compare these nested models, where ΔCFI <0.01 and ΔRMSEA <0.015 indicate a non-significant change in model fit (Cheung & Rensvold, 2002; Chen 2007). To determine the source of an eventually lack of equivalence, MPLUS modification indices were checked (Cigularov et al., 2013). Given the higher presence of poor sleep hygiene in individuals with emotional problems and disturbed sleep (Peach et al. 2016; Edinger et al. 2017;Gupta et al. 2019), the presence of negative emotional symptoms (i.e., DASS-21 total score) and self-reported sleep problems (i.e., MOS sleep problem index) were used as grouping variable.

## 3. Results

### 3.1. Descriptive statistics

Analyses was carried out on a total of 6276 participants, aged 18–82 years (M = 33.62, SD = 13.45), that voluntarily took part in our validation study. Most participants were female (73,2%). People who completed the survey were predominantly resident in the north of Italy, single/never married and childless. In terms of education, participants who had a high school diploma were mainly represented (47%). Students were approximately 29% of the sample. Table 2 summarizes descriptive statistics for each item of the Sleep Hygiene Index. Items means ranged from 1.25 to 3.04. SHI items also presented non-negligible skewness and kurtosis. Specifically, skewness ranged from 0.09 to 3.09, while kurtosis ranged from -1.3 to 9.9. As can be noted, several items exceed the value of |1| for univariate skewness and kurtosis, suggesting a deviation from univariate normality. The SHI total score reported in the present study was 26.61 (SD = 7.12) ranging from 13 to 61. Table 2 presents descriptive statistics such as scales means, range and standard deviations of all the studied variables.

**Table 1.** SHI items characteristics.

| SHI items | Mean | Standard deviation | Skewness | Kurtosis |
| --- | --- | --- | --- | --- |
| SHI_1 | 1.44 | .76 | 1.9 | 3.51 |
| SHI_2 | 2.50 | 1.09 | .46 | -.48 |
| SHI_3 | 2.25 | 1.02 | .73 | .07 |
| SHI_4 | 1.25 | .67 | 3.09 | 9.90 |
| SHI_5 | 2.33 | 1.24 | .62 | -.63 |
| SHI_6 | 1.74 | 1.20 | 1.55 | 1.20 |
| SHI_7 | 2.89 | 1.43 | .09 | -1.30 |
| SHI_8 | 2.23 | 1.08 | .69 | -.19 |
| SHI_9 | 2.67 | 1.39 | .27 | -1.20 |
| SHI_10 | 1.40 | .91 | 2.48 | 5.65 |
| SHI_11 | 1.32 | .78 | 2.92 | 8.68 |
| SHI_12 | 1.56 | .91 | 1.74 | 2.61 |
| SHI_13 | 3.04 | 1.22 | .10 | -.92 |

**Table 2.**
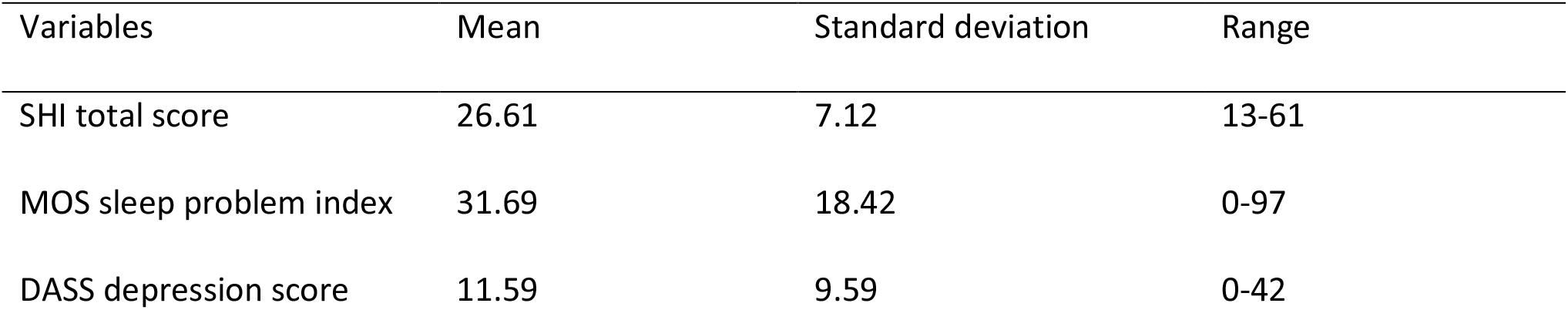
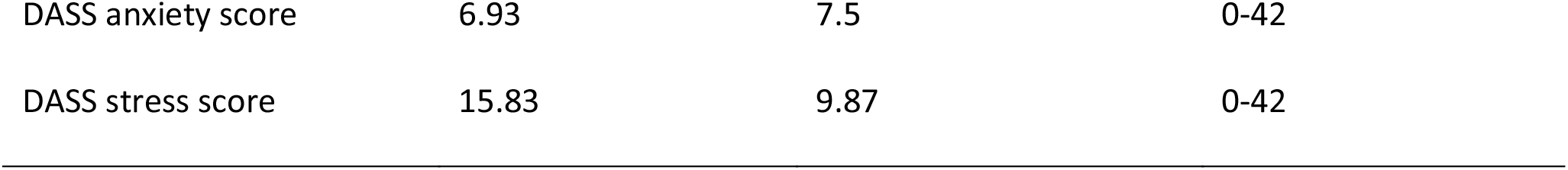
Descriptive statistics of the main scales included in our investigation. These statistics refer to cross-sectional data.

### 3.2. Confirmatory factor analysis

By means of Treiblmaier et al., (2011) procedure, a confirmatory factor analysis was conducted with the robust maximum likelihood method of estimation. The posited sleep hygiene factor was loaded by the four item-composites previously implemented. CFA yielded the following fit indices: χ2=88.72 (df=2, p<0.001); RMSEA=0.083; CFI=0.980; TLI=0.940; SRMR=0.020. All fit indices indicated a good or excellent fit to the data, except for RMSEA that was slightly above the recommended cut-off. However, the RMSEA is strongly influenced by the model degrees of freedom (DFs) showing a worst fit when the DFs are relatively small (Kenny, Kaniskan & McCoach, 2014). For model with small DFs, the RMSEA could suggest an inadequate fit to the data even if the model is correctly specified (Kenny, Kaniskan & McCoach, 2014). Thus, caution with the interpretation of the RMSEA is needed (Taasoobshirazi & Wang, 2016). According to this and considered that our model had only two DFs, we hypothesized that when the DFs will increase during the invariance tests, the RMSEA will be adequate providing an evidence of acceptable fit of the model to the data. Table 3 presents the factor solution. Sleep hygiene construct was significantly loaded by the intended item-composites with all factor loadings greater than 0.55.

**Table 3.** Factor loadings are presented in completely standardized metric. The four indicators are weighted item-composites. All factor loadings are statistically significant (p<0.001).

| SHI factor solution | Standardized factor loadings |
| --- | --- |
| SHI_C1 | 0.625 |
| SHI_C2 | 0.688 |
| SHI_C3 | 0.721 |
| SHI_C4 | 0.566 |

### 3.3. Reliability and validity

The overall reliability of the sleep hygiene dimension was confirmed with a McDonald’s ω coefficient of 0.752. Test-retest reliability at 8-10 months showed an ICC (2,1) of 0.666. In terms of predictive validity, SHI total score significantly correlated with the MOS sleep problem index in cross-sectional analysis (r=0.358; p<.001), and at 8-10 months follow-up (r= 0.410 (p<.001).

Moderate correlations were also found with the DASS depression score (r=0.475; p<.001) and with the DASS anxiety score (r=0.409; p<.001). Finally, SHI was strongly correlated with the DASS stress index (r=0.500; p<.001).

### 3.4. Factorial invariance tests

Factorial invariance tests across people with negative emotional symptoms (i.e., DASS total score; with symptoms n = 1662, without symptoms n = 4615) were performed. The cut-off score was set at the 75th percentile. Configural invariance was reached: χ2=98.74 (df=4, p<0.001); RMSEA=0.087; CFI=0.974; TLI=0.921; SRMR=0.022. Thus, the model was tenable when tested simultaneously on both groups. All fit indices were acceptable, except for the RMSEA that, as explained previously, is affected by the low degrees of freedom (DFs=4). Subsequently, constraints on factor loadings were introduced and metric invariance was obtained (ΔCFI = 0.001; RMSEA improved). As hypothesised, when the degrees of freedom increased, the RMSEA became acceptable (0.067).

Finally, constraints on item intercepts were added and the fit of model decreased in fit (ΔCFI >0.01 ΔRMSEA >0.015). An exploration of the modification indices revealed that an introduced constraint was not tenable (intercepts of item-composites 4). Once the constraint was removed, the level of partial scalar invariance was reached (ΔCFI = 0.001; ΔRMSEA = 0.007) (Byrne, Shavelson & Muthén, 1989). Considering all the results, latent means difference was tested by fixing mean value to zero for the “without symptoms” group (i.e., reference group), while was freely estimated in the “symptoms” group. People with negative emotional symptoms reported higher latent means on the sleep hygiene dimension (0.391; p <.001).

Factorial invariance tests across people with self-reported sleep problems (i.e., MOS sleep problem index; with problematic sleep = 3476, without problematic sleep n = 2801) were also computed. Firstly, configural invariance was obtained with the following fit indices: χ2=86.98 (df=4, p<0.001); RMSEA=0.091; CFI=0.972; TLI=0.915; SRMR=0.024. Again, the RMSEA was potentially affected by the low degrees of freedom. Subsequently, we gradually introduced constraints on factor loadings and intercepts to test the metric and the scalar invariance. Metric invariance was reached with a ΔCFI of 0.005 and, as expected, an improvement of the RMSEA. When constraints of scalar invariance were added, the model showed a significant decrease in fit (ΔCFI >0.01; ΔRMSEA >0.015). Even through the exploration of the modification indices scalar invariance was not feasible, thus latent means comparison was not implemented. All invariance level reached are summarized in Table 4.

**Table 4.**
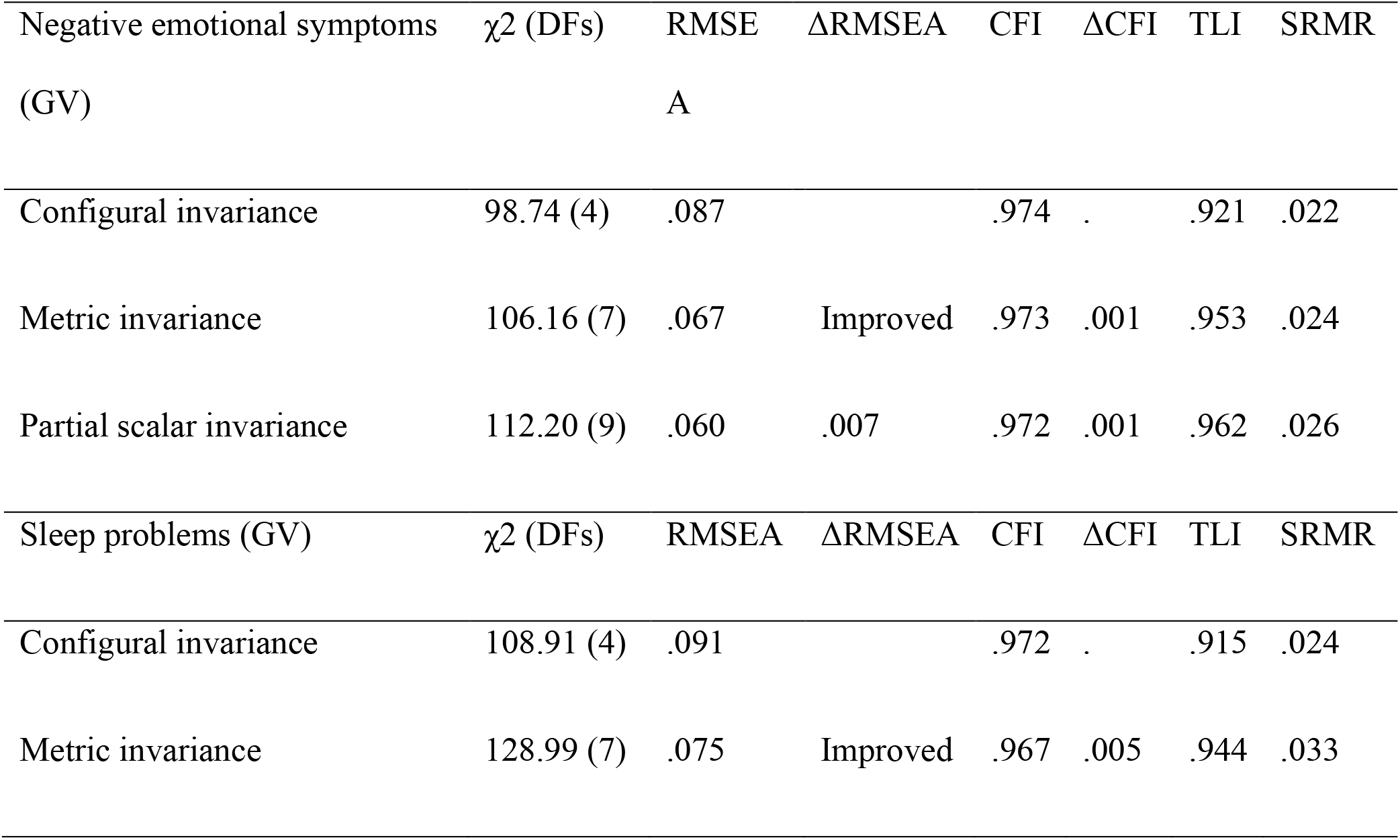
Factorial invariance level reached across negative emotional symptoms and across sleep problems. GV=grouping variable.

## 4. Discussion

The aim of present study was to adapt the SHI into Italian and to examine its psychometric properties for the assessment of sleep hygiene in the Italian general population. Specifically, we evaluated the factor structure, the reliability and the validity of the tool, as well as the factorial invariance of the construct across people with sleep problems groups and across people with emotional distress symptoms.

According to Mastin et coll (2006), the SHI was developed following a formative indicator approach. In a formative model causality flows from indicator to construct (Coltman et al., 2008). Since exploratory factor analysis, confirmatory factor analysis and classic reliability theory assume that causality runs from construct to indicator, a different testing approach was needed. With the aim to overcome identification problem (i.e., a pure formative measurement model cannot be identified by themselves) and to define latent variable only by their antecedent indicator, the Treiblmaier et al., (2011) procedure was implemented. After developing a set of weighted item-composites to take into account the item formative nature, a CFA was carried out. The confirmatory factor analytic findings empirically confirmed the factor model with a satisfactory goodness of fit. All fit indices indicate an adequate fit to the data, except for the RMSEA that was slightly above the recommended cut-off. A possible explanation refers to the nature of this specific fit index. Specifically, RMSEA is strongly influenced by model’s degree of freedom (DFs), indeed it tends to show worst fit when the DFs are relatively small (Breivik & Olsson, 2001). As the DFs decreased, the rejection of models seems to increase when the RMSEA is used (Taasoobshirazi & Wang, 2016). As previously written, for model with small DFs the RMSEA can overstep cut-off falsely suggesting a poor fit even if the model is correctly specified (Kenny, Kaniskan & McCoach, 2014). Given all the above and considering that our model had only two degrees of freedom, we could conclude that a substantial fit of the model to the data was obtained. Results of other fit indices are summarily in line or better compared to other validation studies (e.g., Ozdemir et al., 2015).

Findings also showed an acceptable internal consistency (Omega coefficient=0.752). This result is in line to other validation studies (e.g., Tonon et al., 2020; Ozdemir et al., 2015), whereas is higher than the English version (Mastin et al., 2006), the Chinese version (Tang et al., 2021) and the Nigerian version (Seun-Fadipe et al., 2018) of the tool. In terms of test-retest reliability, an ICC of 0.666 emerged. Considering the time elapsed between the two assessments (i.e., 8-10 months), the SHI seems to have an adequate temporal stability in our sample. For example, the Turkish validation of the tool showed an intraclass correlation coefficients of 0.62 after only a 3-week interval (Ozdemir et al., 2015). These findings demonstrate that our version of the SHI is a reliable checklist for the Italian speaking population. The reliability was also found to be satisfactory and relatively high compared to the other sleep hygiene tools (e.g., SHAPS, Lacks & Rotert, 1986; SHST, Blake & Gòmez, 1998).

In cross-sectional and longitudinal analyses, poor sleep hygiene resulted a significant predictor of poor sleep, supporting the literature on sleep hygiene as a perpetuating factor of sleep disturbances (Morin 1993, Sönmez & Aksoy Derya, 2018; Jung et al. 2019; Booker et al. 2020). Moreover, cross-sectional correlation analyses showed significant associations between sleep hygiene and anxiety, depression, and stress symptoms. Taken together, these results provide evidence for the predictive validity of the SHI. Sleep hygiene and emotional problems are closely related. For example, individuals with depression often report poor sleep hygiene (Gupta et al. 2019), and results of experimental researched showed that the implementation of sleep hygiene practices may improve depression (Rahimin et al. 2016). To the best of our knowledge, no studies have investigated the factorial invariance of the construct according to clinical variables. In order to fill this gap, factorial invariance test across people with emotional distress (i.e., group with significative presence of emotional distress symptoms vs group without emotional distress symptoms) and across people with sleep problems (i.e., group with self-reported sleep problems vs group without self-reported sleep problems) were implemented. Findings showed that configural invariance was achieved in both tests. Thus, we could conclude that the basic organization of the construct was the same according to the mentioned grouping variables. Metric invariance was also reached for both tests. Hence, each weighted item-composites seems to contribute similar to latent construct across mentioned groups. Lastly, a partial scalar invariance was obtained only when considered the presence of emotional distress symptoms as grouping variable. When latent mean comparison was implemented, participants with a significative presence of negative emotional symptoms reported higher latent means on the sleep hygiene dimension. This result is consistent with previous literature on sleep hygiene in clinical samples with emotional disorders (Peltz & Rogge, 2016) and informs about the robustness of the SHI in symptomatic samples.

The present study encompasses several limitations that need to be highlighted. First, a convenience sample was used, and this could affect the generalization of the survey results to the population as a whole. Second, while longitudinal analysis may allow a hypothesise causal impact of sleep hygiene on sleep symptoms, causal relations between sleep hygiene and mental health symptoms cannot be inferred due to the cross-sectional nature of these data. Thus, future longitudinal studies are needed to examine the causal associations between sleep hygiene and mental health variables. Moreover, considering the relationship between sleep hygiene and psychopathological symptoms (e.g., Peltz & Rogge, 2016; Gupta et al. 2019), future replication studies should be conducted to prove the adequacy of the instrument on large clinical samples. Finally, an alternative sleep hygiene measure will be required to provide the convergent validity of the SHI as well as a measure to assess the divergent validity.

### 4.1. Conclusions

Our result support the validity and reliability of the Italian version of the SHI in assessing adherence to sleep hygiene practices in the general population. Importantly, the SHI showed robust psychometric properties both in healthy individuals and in individuals reporting mental health symptoms. Moreover, it also showed to predict sleep quality after 8-10 months. Thus, it is advisable to use this version of the SHI in both research and clinical practice.

## Data Availability

The datasets presented in this article are not readily avaible. Requests to access the datasets should be directed to

## References

American Academy of Sleep Medicine (2014). International classification of sleep disorders: Diagnostic and coding manual. 3 ed. Westchester, Illinois: American Academy of Sleep Medicine.

Bagozzi, R.P., Yi, Y. (2012). Specification, evaluation and interpretation of structural equation models. Journal of the Academy of Marketing Science, 40(1), 8–34.

Blake, D.D., Gomez M.H. (1998). A scale for assessing sleep hygiene: preliminary data. Psychol Rep, 83(3 Pt 2):1175–1178.

Bollen, K.A., Diamantopoulos, A. (2017). In defense of causal-formative indicators: A minority report. Psychological methods, 22(3), 581–596.

Bollen, K.A. (1989). A new incremental fit index for general structural equation models.

Booker, L.A., Barnes, M., Alvaro, P., Collins, A., Chai-Coetzer, C.L., McMahon, M., Lockley, S.W., Rajaratnam, S.M.W., Howard, M.E., Sletten, T.L. (2020). The role of sleep hygiene in the risk of Shift Work Disorder in nurses. Sleep, 42(2).

Bottesi, G., Ghisi, M., Altoè, G., Conforti, E., Melli, G., Sica, C. (2015). The Italian version of the Depression Anxiety Stress Scales-21: Factor structure and psychometric properties on community and clinical samples. Comprehensive psychiatry, 60, 170–181.

Breivik, E., Olsson, U.H. (2001). Adding Variables to Improve Fit: The Effect of Model Size on Fit Assessment in LISREL.’’ Pp. 169–94 in Structural Equation Modeling: Present and Future, edited by Robert Cudeck, S. H. C. du Toit, and Dag Sö rbom. Lincolnwood, IL: Scientific Software International

Byrne, B.M., Shavelson, R.J., Muthén, B. (1989). Testing for the equivalence of factor covariance and mean structures: the issue of partial measurement invariance. Psychological bulletin, 105 (3)

Chen, F.F. (2007). Sensitivity of goodness of fit indexes to lack of measurement invariance. Structural Equation Modeling, 14:464–504.

Cheung, G.W., Rensvold, R.B. (2002). Evaluating goodness-of-fit indexes for testing measurement invariance. Structural Equation Modeling, 9(2), 233–255.

Cigularov, K. P., Lancaster, P. G., Chen, P. Y., Gittleman, J., Haile, E. (2013). Measurement equivalence of a safety climate measure among Hispanic and White Non-Hispanic construction workers. Safety Science, 54, 58–68.

Cohen, J. (1988). Statistical power analysis for the behavioral sciences.

Erlbaum Coltman, T., Devinney, T. M., Midgley, D. F., Venaik, S. (2008). Formative versus reflective measurement models: Two applications of formative measurement. Journal of Business Research, 61(12), 1250–1262.

Costa, J., Helou, S., Sleilaty, G., Costa T., El Helou J. (2021). Validity and reliability of an Arabic version of the Sleep Hygiene Index. Sleep Med, 80:260–264.

Edinger, J.D., Means, M.K., Carney, C.E., Manber, R. (2017). Psychological and Behavioral Treatments for Insomnia II: Implementation and Specific Populations. In: Principles and Practice of Sleep Medicine (Sixth Edition).

Hays, R.D., Stewart, A.L., (1992). Sleep measures. In: Stewart AL, Ware JE, editors. Measuring functioning and well-being: the Medical Outcomes Study approach. Durham (NC): Duke University Press, 235–259.

Hays, R. D., Martin, S. A., Sesti, A. M., Spritzer, K. L. (2005). Psychometric properties of the Medical Outcomes Study Sleep measure. Sleep medicine, 6(1), 41–44.

Hermosilla, I.T., Alvarado, J. (2016). Best Alternatives to Cronbach’s Alpha Reliability in Realistic Conditions: Congeneric and Asymmetrical Measurements. Frontiers in Psychology. 7.

Irish, L.A., Kline, C.E., Gunn, H.E., Buysse, D.J., Hall, M.H. (2015). The role of sleep hygiene in promoting public health: A review of empirical evidence. Sleep Med Rev, 22, 23–36.

Jung, S.Y., Kim, H.S., Min, J.Y., Hwang, K.J., Kim, S.W. (2019). Sleep hygiene-related conditions in patients with mild to moderate obstructive sleep apnea. Auris Nasus Larynx. 2019 Feb;46(1):95–100.

Kenny, D.A., Kaniskan, B., McCoach, D.B. (2014). The Performance of RMSEA in Models With Small Degrees of Freedom. Sociological Methods & Research, 44(3), 486–507.

Kleitman, N. (1939). Sleep and wakefulness. Univ. Chicago Press.

Lacks, P., Rotert, M. (1986). Knowledge and practice of sleep hygiene techniques in insomniacs and good sleepers. Behav Res Ther, 24(3):365–368.

Little, T.D., Rhemtulla, M., Gibson, K., Schoemann, A.M. (2013). Why the items versus parcels controversy needn’t be one. Psychological Methods, 18(3), 285–300.

Lovibond, P.F., Lovibond, S.H. (1995). The structure of negative emotional states: Comparison of the Depression Anxiety Stress Scales (DASS) with the Beck Depression and Anxiety Inventories. Behaviour Research and Therapy, 33, 335–343.

Hu, L., Bentler, P.M. (1999). Cutoff criteria for fit indexes in covariance structure analysis: conventional criteria versus new alternatives. Struct Equ Model Multidiscip J 6(1):1–55.

MacCallum R.C., Browne M.W. (1993), The Use of Causal Indicators in Covariance Structure Models: Some Practical Issues. Psychological Bulletin, 114 (3), 533–41.

Mastin, D.F., Bryson, J., Corwyn, R. (2006). Assessment of sleep hygiene using the Sleep Hygiene Index. Journal of behavioral medicine, 29(3), 223–227.

McDonald R. P. (1999). Test theory: A unified treatment. Mahwah, NJ: Lawrence Erlbaum.

Meredith, W. (1993). Measurement invariance, factor analysis and factorial invariance. Psychometrika, 58(4), 525–543.

Ozdemir, P.G., Boysan, M., Selvi, Y., Yildirim, A., Yilmaz, E. (2015). Psychometric properties of the Turkish version of the Sleep Hygiene Index in clinical and non-clinical samples. Comprehensive psychiatry, 59, 135–140.

Palagini, L., & Manni, R. (2018). Misurare il sonno. Repertorio delle scale di valutazione dei disturbi del sonno. Minerva medica: 2017.

Peltz, J.S., Rogge, R.D. (2016). The indirect effects of sleep hygiene and environmental factors on depressive symptoms in college students. Sleep Health, 2(2), 159–166.

Prados, G., Chouchou, F., Carrión Pantoja, S., Fernández Puerta, L., Pérez Mármol, J.M. (2021). Psychometric properties of the Spanish version of the Sleep Hygiene Index. Research in Nursing & Health, 44(2), 393–402.

Rahimi, A., Ahmadpanah, M., Shamsaei, F., Fatemeh, C., Bahmani, D.S., Holsboer-Trachsler, E., Brand, S. (2016). Effect of adjuvant sleep hygiene psychoeducation and lorazepam on depression and sleep quality in patients with major depressive disorders: results from a randomized three-arm intervention. Neuropsychiatric Disease and Treatment, 12, 1507–1515.

Seun-Fadipe, C.T., Aloba, O.O., Oginni, O.A., Mosaku, K.S. (2018). Sleep hygiene index: psychometric characteristics and usefulness as a screening tool in a sample of Nigerian undergraduate students. Journal of Clinical Sleep Medicine, 14(8), 1285–1292.

Shrout, P.E., Fleiss, J.L. (1979). Intraclass correlations: uses in assessing rater reliability. Psychol Bull, 86:420–428.

Sönmez, A., Aksoy Derya, Y. (2018). Effects of sleep hygiene training given to pregnant women with restless leg syndrome on their sleep quality. Sleep Breath, 22(2):527–535.

Spielman, A., Caruso, L., Glovinsky P. (1987). A behavioral perspective on insomnia treatment. Psychiatr Clin North Am, 10:541–553.

Taasoobshirazi, G., Wang, S. (2016). The performance of the SRMR, RMSEA, CFI, and TLI: An examination of sample size, path size, and degrees of freedom. Journal of Applied Quantitative Methods, 11(3), 31–39.

Tabachnick, B.G., Fidell, L.S. (2007). Using Multivariate Statistics. Pearson/Allyn & Bacon.

Tanaka, J.S (1993). Multifaceted conceptions of fit. In: Kennet AB, Long JS, editors. Structural equation models.

Tang, Z., Li, X., Zhang, Y., Li, X., Zhang, X., Hu, M., & Wang, J. (2021). Psychometric analysis of a Chinese version of the Sleep Hygiene Index in nursing students in China: a cross-sectional study. Sleep Medicine, 81, 253–260.

Tonon, A.C., Amando, G.R., Carissimi, A., Freitas, J.J., Xavier, N.B., Caumo, G. H., … & Hidalgo, M. P. (2020). The Brazilian-Portuguese version of the Sleep Hygiene Index (SHI): validity, reliability and association with depressive symptoms and sleep-related outcomes. Sleep Science, 13(1), 37.

Treiblmaier, H., Bentler, P. M., & Mair, P. (2011). Formative constructs implemented via common factors. Structural Equation Modeling, 18(1), 1–17.

